# “Is this Herpes or Syphilis?”: Latent Dirichlet Allocation Analysis of Sexually Transmitted Disease-Related Reddit Posts During the COVID-19 Pandemic

**DOI:** 10.1101/2022.02.13.22270890

**Authors:** Amy K Johnson, Runa Bhaumik, Debarghya Nandi, Abhishikta Roy, Supriya D Mehta

## Abstract

**Background:** Sexually Transmitted Diseases (STDs) are common and costly, impacting approximately one in five people annually. Reddit, the sixth most used internet site in the world, is a user-generated social media discussion platform that may be useful in monitoring discussion about STD symptoms and exposure.

**Objective:** This study sought to define and identify patterns and insights into STD related discussions on Reddit over the course of the COVID-19 pandemic.

**Methods:** We extracted posts from Reddit from March 2019 through July 2021. We used a machine learning text mining method, Latent Dirichlet Allocation (LDA), to conduct a text analysis to identify the most common topics discussed in the Reddit posts. We then used word clouds, qualitative topic labelling, and spline regression to characterize the content and distribution of topics observed.

**Results:** Our extraction resulted in 24,311 total posts. LDA Coding showed that with 8 topics for each time period we achieved high coherence values (pre-COVID=0.41, pre-vaccine=0.42; post-vaccine=0.44). While most topic categories remained the same over time, the relative proportion of topics changed and new topics emerged. Spline regression revealed some key terms had variability in the percentage of posts that coincided with COVID-19 pre- and post-periods, while others were uniform across the study periods.

**Conclusions:** Our study’s use of Reddit is a novel way to gain insights into STD symptoms experienced, potential exposures, testing decisions, common questions, and behavior patterns (e.g., during lock down periods). For example, reduction in STD screening may result in observed negative health outcomes due to missed cases, which also impacts onward transmission. As Reddit use is anonymous, users may discuss sensitive topics with greater detail, and more freely than in clinical encounters. Data from anonymous Reddit posts may be leveraged to enhance understanding of the distribution of disease and need for targeted outreach/screening programs. This study demonstrates Reddit has feasibility and utility to enhance understanding of sexual behaviors, STD experiences, and needed health engagement with the public.

## Introduction

More than 2.5 million cases of chlamydia, gonorrhea, and syphilis were reported in 2019 with sexually transmitted diseases (STD) cases reaching an all-time high for the 6^th^ consecutive year in the U.S. [1] STDs are common and costly, impacting approximately one in five people annually and accounting for $16 billion in annual healthcare costs.[2] New data from the Centers for Disease Control and Prevention (CDC) demonstrate that during the start of the COVID-19 pandemic (March-April 2020), reported STD cases dramatically decreased compared to the same time in 2019. At that point the current cumulative totals for STD cases compared to 2019 were 1% lower for primary and secondary syphilis, 7% lower for gonorrhea, and 14% lower for chlamydia.^[3]^ While case reports were lower for the first part of 2020, later in the year cases rebounded and were on track to surpass 2019 totals.[3]

Multiple factors likely contributed to the observed decrease in reported STD cases during the early phases of the COVID-19 pandemic. Due to clinic restrictions of in-person visits reduced screening for asymptomatic patients occurred. The CDC provided guidance for sexual health services to prioritize patients based on symptoms and risk, with delaying routine screening until after the emergency response.[4] Many health department staff were re-deployed from STD tracking to COVID-19 contact tracing and control.[5] Fifty-seven percent of disease intervention specialists reported that they were reassigned from STD to COVID-19 services, limiting the workforce available to provide STD prevention, screening, and treatment. [5] Finally, national stay-at-home orders were issued during phases of the pandemic that were designed to reduce the spread of COVID-19 but may also have reduced STD transmission by reducing sexual behavior outside of the household, limiting the number of new sexual partners, and restricting sexual networks.[6]

Recent estimates indicate that 80% of all Internet users report accessing health information online.[7] As the Internet can be accessed anonymously and at any time, users can seek STD information and resources confidentially which may facilitate more frequent and open disclosure of symptoms and exposure experiences.[8] Reddit, the sixth most used internet site in the world, is a user-generated social media discussion platform that may be useful in monitoring discussion about STD symptoms and exposure.[9] Topic specific Reddit discussions (sub-reddits) dedicated to discussing sexual health and STDs may provide valuable insight to exposure, symptoms, testing, and sexual behavior during the COVID-19 pandemic. However, to derive meaningful and replicable information, the complexity of high-volume text data needs numerical structure implemented in an unbiased way. Latent Dirichlet allocation (LDA) is a natural language processing method that identifies common words and topics in text and allows experts to assess common themes among findings.[10] This study sought to define and identify patterns and insights into STD related discussions on Reddit via LDA over the course of the COVID-19 pandemic.

## Methods

### Institutional Review Board Approval

The study protocol was determined to be non-human subjects research by the Ann & Robert H. Lurie Children’s Hospital Institutional Review Board (IRB#2022-4964) because of the use of publicly available non-identifiable data.

### Data Extraction

The current study used publicly available data from the online discussion forum, Reddit. Reddit is an anonymous social media site that is user-generated and discussion based. The site is organized into “sub-reddits” that are content specific. Posts were extracted from two subreddits: “STD” and “sexual health” (r/STD; r/sexualhealth). The pushshift.io Reddit API was used for searching Reddit comments and submissions.[11] Reddit’s official API (Reddit 2021) was used to collect posts and associated metadata (date) from r/STD and r/sexualhealth from March 2019 to July 2021 resulting in 24,311 posts.[10] Only English posts were included in the analysis. Figure 1 displays the number of posts that were extracted from each subreddit for the timeframes used in analysis. Pre-COVID was defined as March 2019 through February 2020 (8421 posts), COVID/pre-vaccination as April 2020 to December 2020 (8169 posts), COVID/post-vaccination from January 2021 to August 2021 (6908 posts) and an inflection period as March 2020 (813 posts).

**Figure 1:**
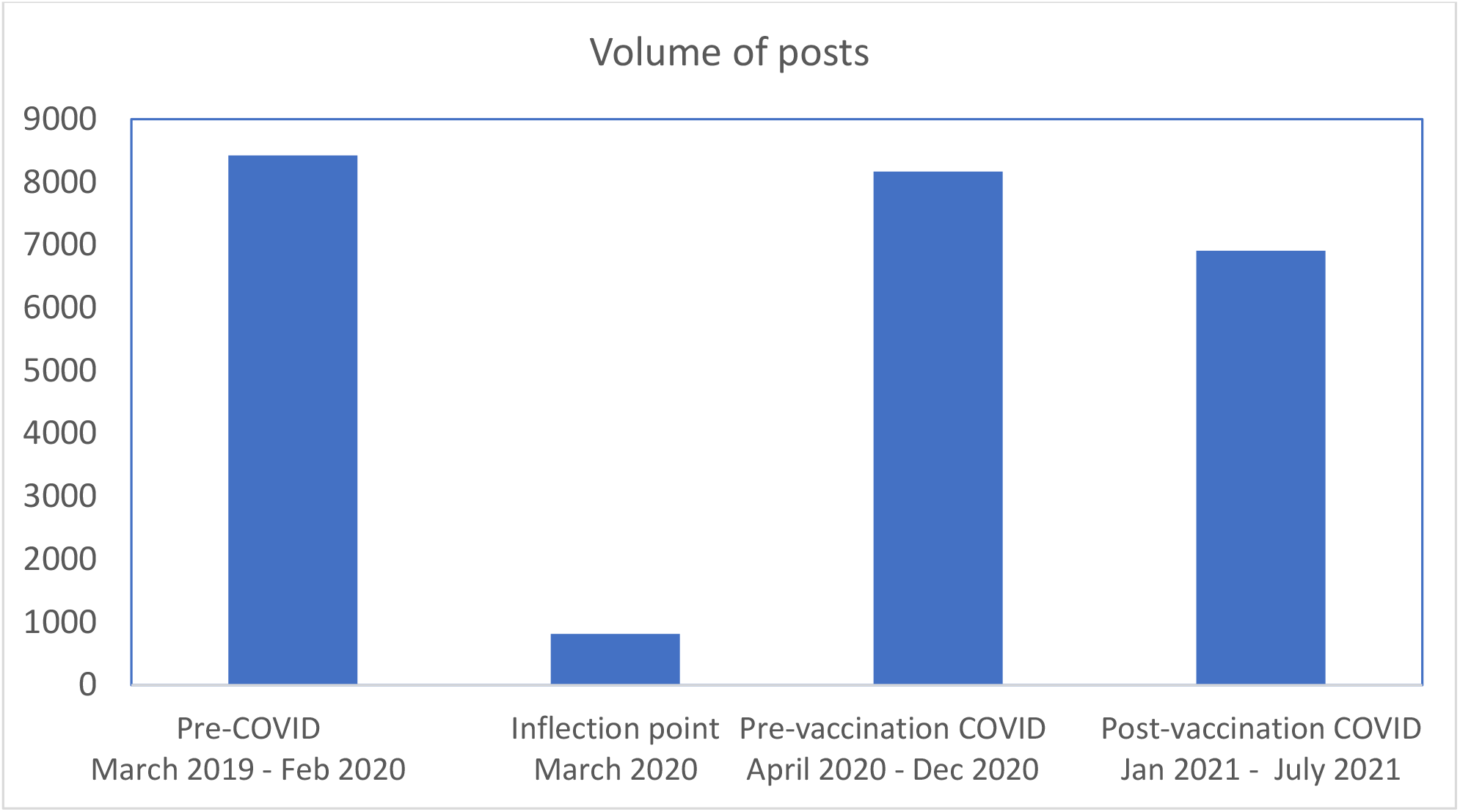
Distribution of volume of posts for study time periods from February 2019 to August 2021 resulting in a total of 24,311 posts

#### Data preprocessing

Data preprocessing steps were conducted following common approaches in natural language processing (NLP).[12] Preprocessing eliminates some of the inconsistencies in the data and reduces the content to useable text. Four preprocessing steps were completed on each line from the text file to extract and clean each title, body, and comment separately: (1) removal of URLs; (2) tokenization; (3) punctuation and stop word removal; and (4) lemmatization.[13-15]

#### Statistical Analysis

##### Latent Dirichlet Allocation

We utilized an increasingly popular machine learning text mining method, Latent Dirichlet Allocation (LDA), to conduct a text analysis identifying the most common topics discussed in the Reddit posts.[16] LDA is a statistical generative model that discovers latent semantic topics in large collections of text documents (posts in our study) where each document results from random mixtures over latent topics and each topic is characterized by a distribution over words. The model is presented in plate notation in Figure 2.[10] Both the topics and words have a Dirichlet prior distribution, respectively, with α being the parameter of the per-document Dirichlet prior on the topics, and β being the parameter of the per-word Dirichlet prior on the words. θ_m_ is the topic distribution for document m. ϕ_k_ is the word distribution for topic k. Z_nm_ is the topic for the n_th_ word in the m_th_ document. W_nm_ is the actual n_th_ word in the m_th_ document. Considering the nature of its structure, LDA is a multiple-level hierarchical Bayesian model.

**Figure 2:**
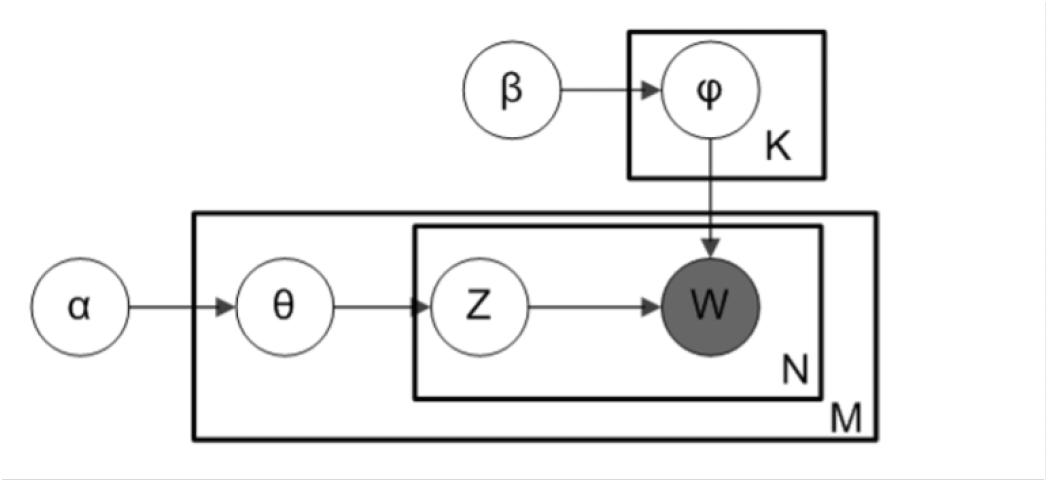
Latent Dirichlet Allocation (LDA) in plate notation

**Figure 3:**
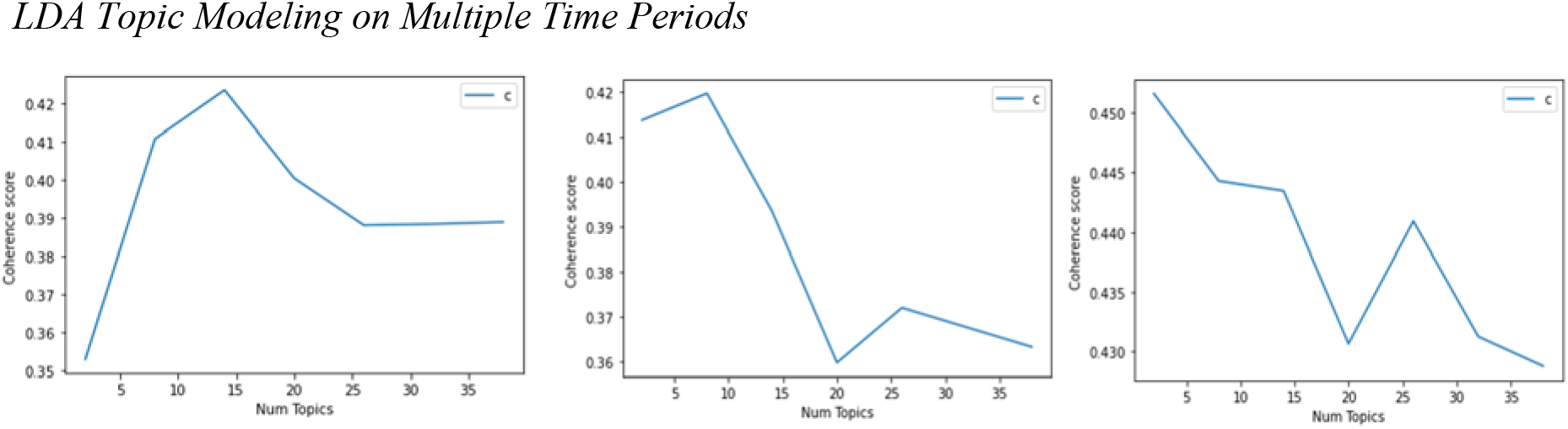
Left: pre-COVID Middle: pre-vaccine Right: post-vaccine *Visualizations*

To conduct the LDA, we converted the corpus to a document-term matrix, comprising rows representing original posts and columns representing each word in the corpus. Each cell in the document-term-matrix contains the frequency of times a specific word (defined by the column) occurred in a specific post (defined by the row). From this document-term-matrix, the entire corpus was represented, including patterns of words that commonly occur together within the same post. We used the *gensim* library to perform LDA model estimation, which determined sets of words that appeared frequently together in posts across sexual health subreddits.[13]

The LDA model then outputs a topic-document matrix, representing the relative importance of each topic in each document. Models were applied to pre-covid posts spanning March 2019 through February 2020 (8421 posts), pre-vaccination posts ranging from April 2020 to December 2020 (8169 posts) and post-vaccination posts between January 2021 to August 2021(6908 posts). For topic modeling we excluded posts for inflection period (March 2020, 813 posts).

A key process in LDA is to estimate the optimal number of topics. To estimate the number of topics, we used topic coherence index, which is the most consistent measure of human interpretability.[17] Topic Coherence measures score a single topic by measuring the degree of semantic similarity between high scoring words in the topic. These measurements help distinguish between topics that are semantically interpretable topics and topics that are artifacts of statistical inference. The higher the topic coherence score, the better the quality of the model. To avoid overfit and sparsity and improve inference, we selected the number of topics as 8. Topics were reviewed and labelled independently by two experts in STD epidemiology and control (AKJ, SDM). Once independent review was completed labels were discussed until consensus was reached, with 100% agreement resulting.

#### WordCloud

A word cloud is a text visualization technique that focuses on the frequency of words and correlates the size and opacity of a word to its frequency within a text body. The output is usually an image that depicts different words in different sizes and opacities relative to the word frequency. Separate frames were created for posts containing the following terms *Chlamydia, Gonorrhea*, and *Syphilis, Gonorrhea/Discharge/Dysuria, and Syphilis/Chancre/Ulcer*. After data preprocessing was complete, each string was passed to the WordCloud function in Python to generate a wordcloud.[18] For WordCloud visualization, we chose three etiologic terms (chlamydia, gonorrhea, syphilis) and three of the most common terminologies from topic search: herpes/HSV/HPV (as a single topic, due to correlations), diagnosis/testing, STI/STD. Each separate wordcloud was formed by searching on each word in the topic.

#### Splined regression plots

To identify patterns in change of proportion of posts related to certain search terms relative to the total number of posts in a particular month over the entire study period (spanning from March 2019 to June 2021), a spline plot was created. The Pre-COVID, Inflection Period, Pre-Vaccination COVID, and Post-Vaccination COVID periods are highlighted on the plots for better understanding of search trends across time. The plots were created using *ggplot2* package in R.[19] For spline regression, we used cubic B-spline basis with two boundary knots and one interior knot placed at the median of the observed data values. As with the wordclouds, we created three plots based on etiology (chlamydia, gonorrhea, and syphilis), and three plots based on common topics (diagnosis/test/tested, herpes/HSV, gonorrhea/dysuria/discharge).

## Results

### Reddit Posts

Out of 24,311 total posts, the average monthly posts were 701.75 per month during the pre-COVID period, 907.67 during the COVID pre-vaccine period, 863.50 covid-post vaccine, but with substantial variability from month-to-month and within each time period. The average monthly posts per period demonstrate growth in the sub-reddits volume during COVID. Figure 4 displays the number of posts per month by observation period. May 2019 consisted of 210 posts and August 2021 of 169 posts, two of the lowest volumes recorded, both of which were preceded by two months of high-volume posts.

**Figure 4.**
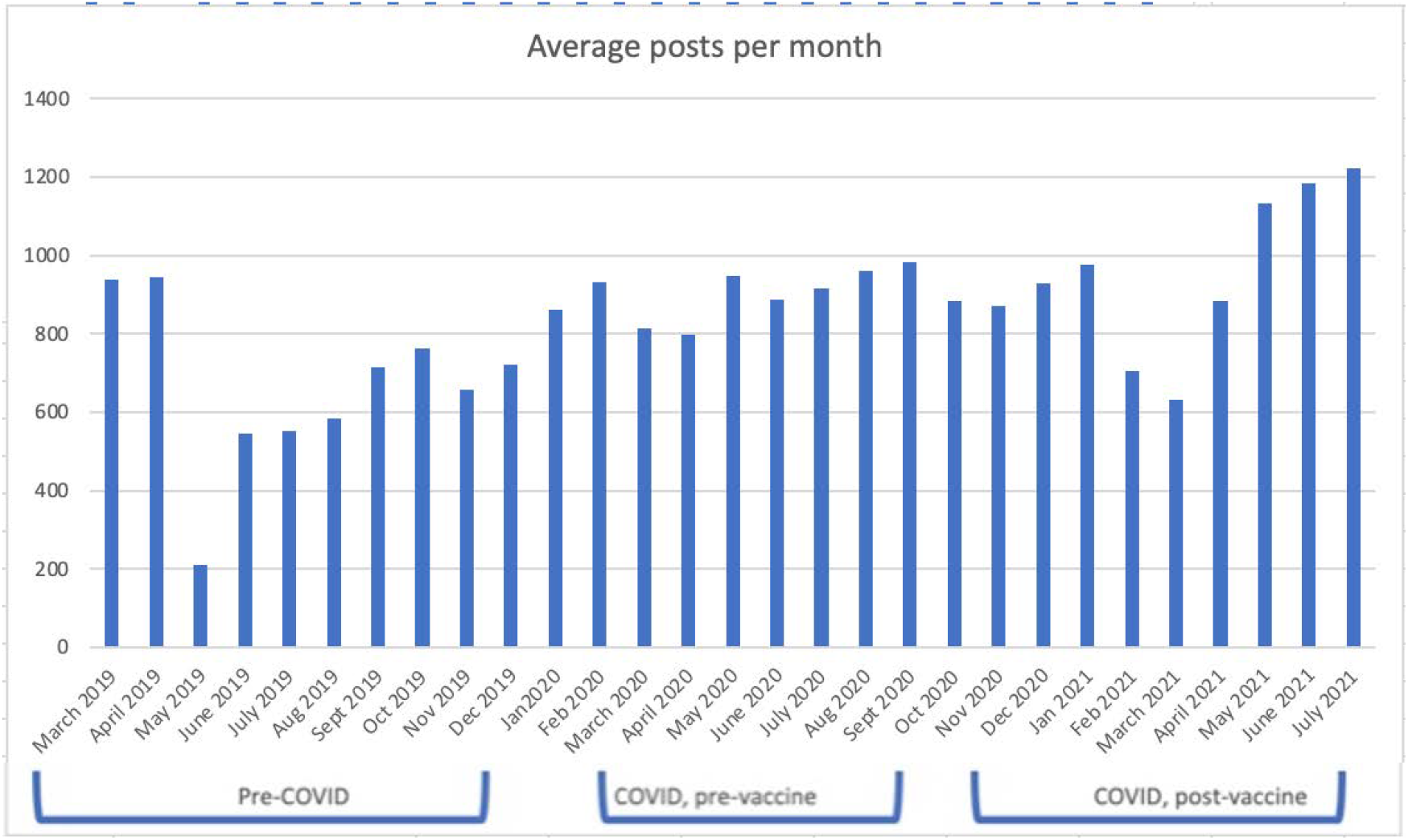
Number of posts per month, by period

LDA Coding showed that with 8 topics for each time period we achieved high coherence values (pre-COVID=0.41, pre-vaccine=0.42; post-vaccine=0.44). Figure 5 shows the distribution of topic posts in pre-COVID, pre-vaccination and post-vaccination “STD” and “sexual health” subreddits over the 8 topics extracted using LDA. While most topic categories remained the same over time, the relative proportion of topics changed and new topics emerged. In the pre-COVID period a general category of “STD Risk” emerged with no specific etiology or mention of symptoms with words such as “negative” and “exposure” in the top 10 terms associated with the topic. “HPV” and “warts” as terms do not appear in the pre-COVID period. There was specific language surrounding herpes symptoms (e.g., “outbreak”) and diagnosis (e.g., testing, and positive or negative) and introduction of “HSV” in the post-vaccination period whereas words used in conjunction with herpes in previous periods were primarily related to images and non-specific symptoms (e.g., “redness, bumps”). Moreover, while the “herpes image” topic category included non-specific symptoms (e.g., “bump”, “redness”), this diverged during COVID periods, with a topic category emerging for penile “bump” without mention of herpes. In the post-vaccination period, the “oral sex/STD question” topic includes the term “penis”, while the topic exists in the other two periods it does not include “penis” as a top 10 term.

**Figure 5:**
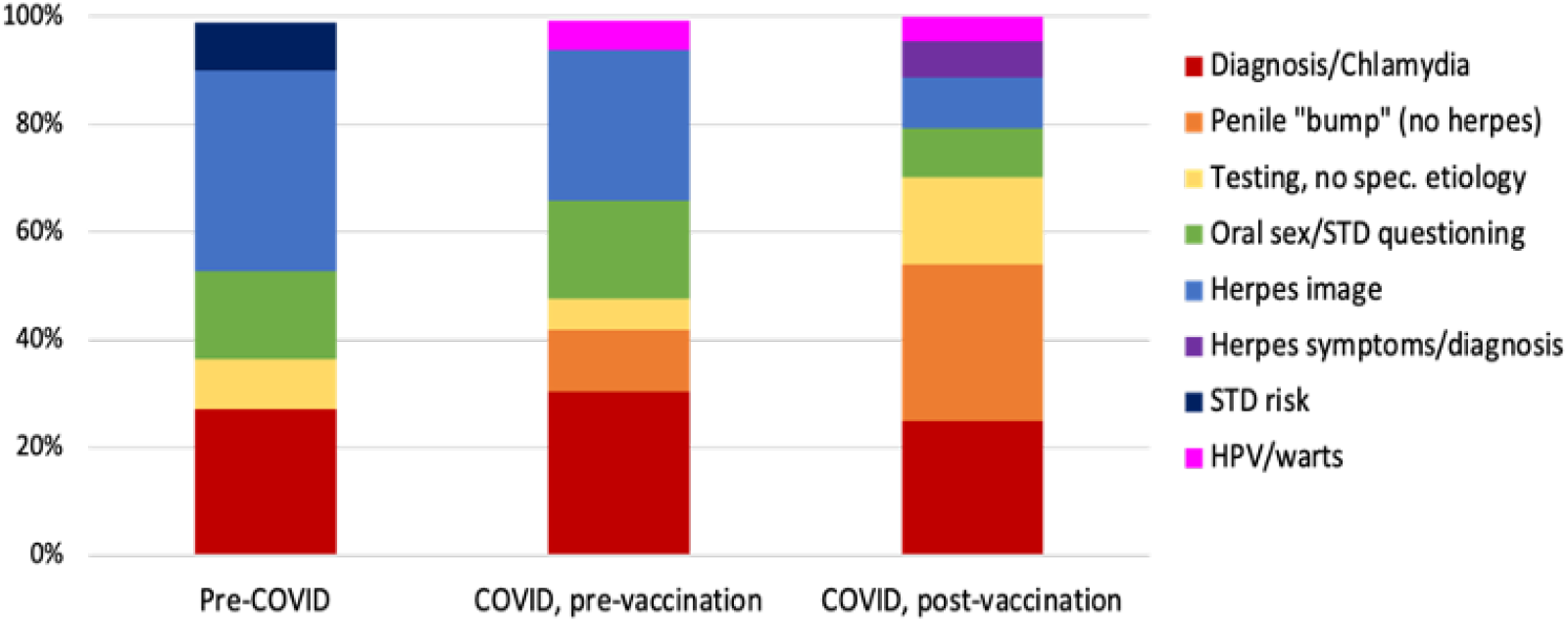
Distribution of posts: the proportion of documents that are assigned to each topic.

**Table 1.**
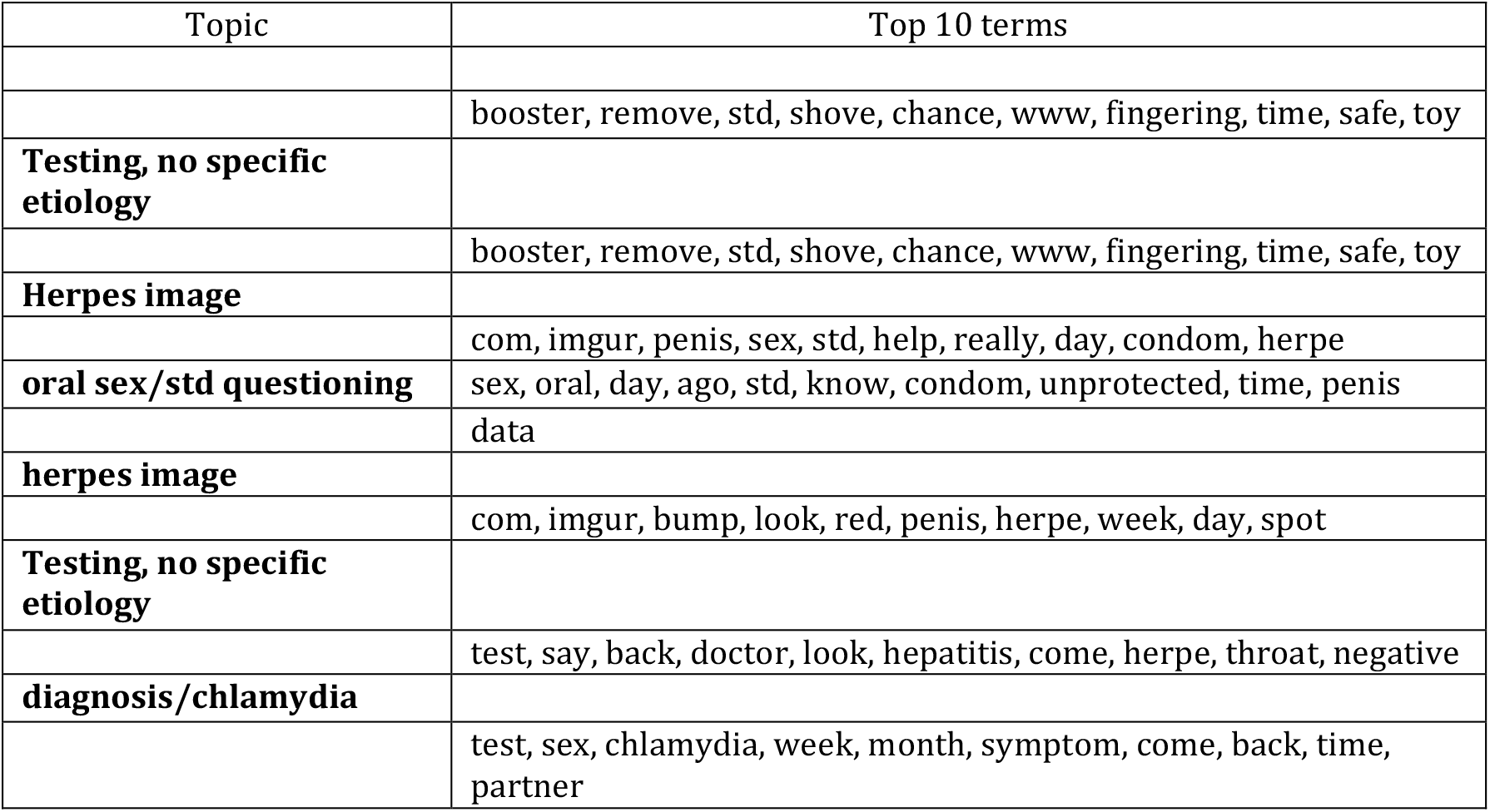
Pre-COVID topic and the 10 terms derived from an LDA model created on three different time periods: pre-COVID, pre-vaccination, and post-vaccination

| Topic | Top 10 terms |
| --- | --- |
|  | booster, remove, std, shove, chance, www, fingering, time, safe, toy |
| **Testing, no specific etiology** |  |
|  | booster, remove, std, shove, chance, www, fingering, time, safe, toy |
| **Herpes image** |  |
|  | com, imgur, penis, sex, std, help, really, day, condom, herpe |
| **oral sex/std questioning** | sex, oral, day, ago, std, know, condom, unprotected, time, penis |
|  | data |
| **herpes image** |  |
|  | com, imgur, bump, look, red, penis, herpe, week, day, spot |
| **Testing, no specific etiology** |  |
|  | test, say, back, doctor, look, hepatitis, come, herpe, throat, negative |
| **diagnosis/chlamydia** |  |
|  | test, sex, chlamydia, week, month, symptom, come, back, time, partner |

**Table 2.**
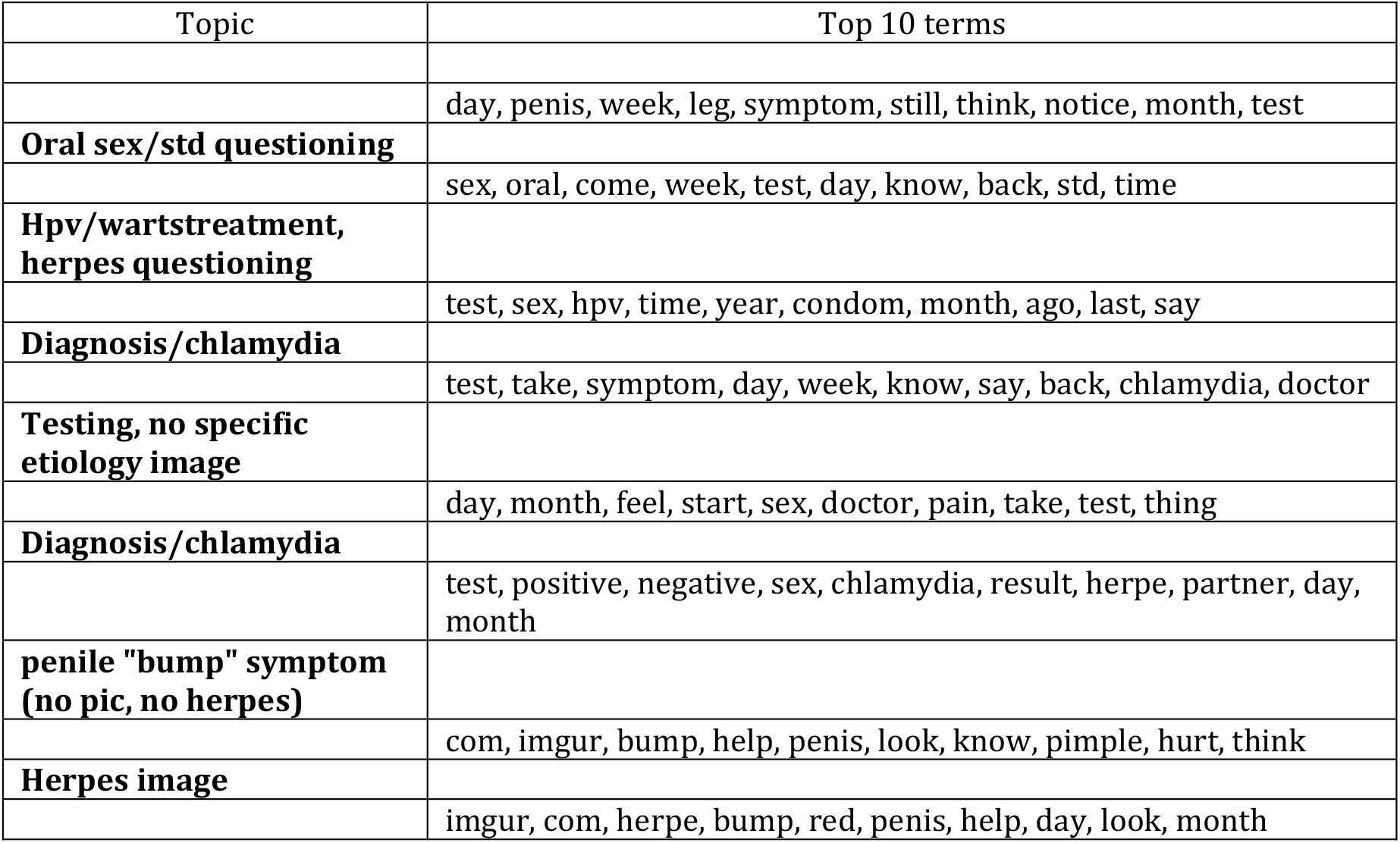
Pre-vaccination topic and the 10 terms derived from an LDA model created on three different time periods: pre-COVID, pre-vaccination, and post-vaccination

**Table 3.** Post-vaccination topic and the 10 terms derived from an LDA model created on three different time periods: pre-COVID, pre-vaccination, and post-vaccination

| Topic | Top 10 terms |
| --- | --- |
| **herpes symptoms/diagnosis** |  |
|  | test, herpe, hsv, sex, know, outbreak, negative, positive, genital, risk |
| **herpes image** |  |
|  | com, imgur, herpe, sex, look, help, remove, know, oral, bump |
| **hpv/warts treatment, herpes questioning** |  |
|  | wart, herpe, ibb_co, com, www_reddit, comment, remove, hpv, month, skin |
| **diagnosis/chlamydia** |  |
|  | day, month, feel, start, sex, doctor, pain, take, test, thing |
| **Testing, no specific etiology** |  |
|  | test, remove, std, day, sex, week, negative, help, time, oral |
| **penile "bump" symptom (no pic, no herpes)** |  |
|  | penis, bump, sex, day, know, std, feel, look, condom, time |
| **oral sex/std questioning** |  |
|  | sex, know, test, week, say, think, time, symptom, oral, tell |
| **penile "bump" symptom** |  |
|  | bump, com, look, imgur, week, ago, day, penis, red, notice |

#### Word Clouds

While the terms in the above listed topic models are informative, we used WordCloud visualizations to better understand the relative importance of these words within each topic based on etiology and general terms over the study period. The terms that appear larger appear more frequently within the topic, with the terms in smaller font appearing less frequently. Figure 6a-f displays the wordclouds for six specific topics, for example figure 6e displays terms clustered with Herpes/HSV/HPV such as “imgur” (denoting a picture uploaded), “bump”, “pain”, and “outbreak”.

**Figure 6a-f.**
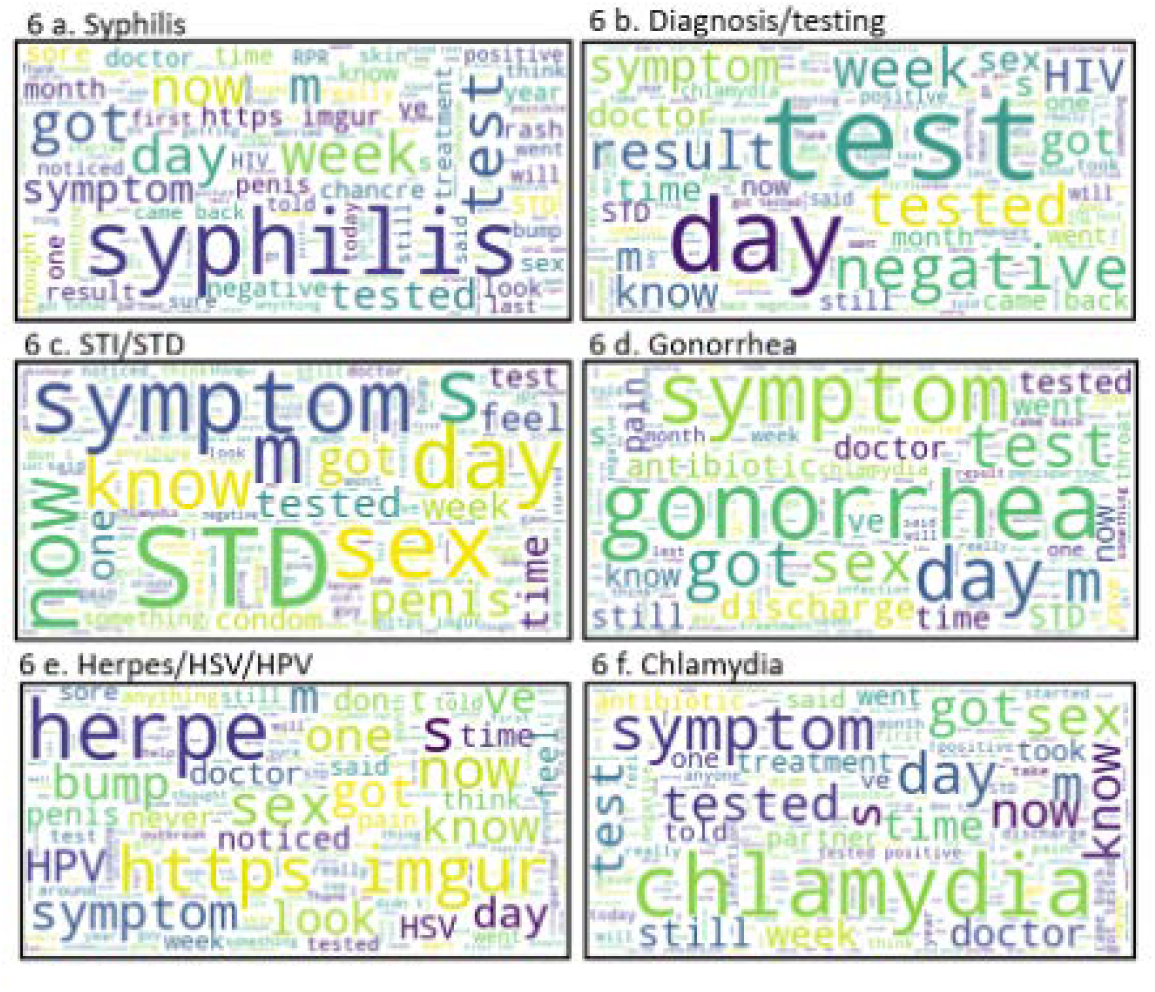
Word cloud by key term

#### Splined regressions

As shown in the series of splined regressions in Figure 7a-g there is some variability in the percentage of posts by key terms over the study periods. While some regressions are “flat” (i.e., uniform) across the study periods, others display variability that coincides with the COVID periods. For example, 7g displays the regression for posts with key terms “diagnose/test/tested”, there is some variability in the percentage of posts in the different COVID periods with pre-COVID and post-vaccine COVID volume being similar to each other and pre-vaccine COVID having a lower percentage of posts.

**Figure 7a-g.**
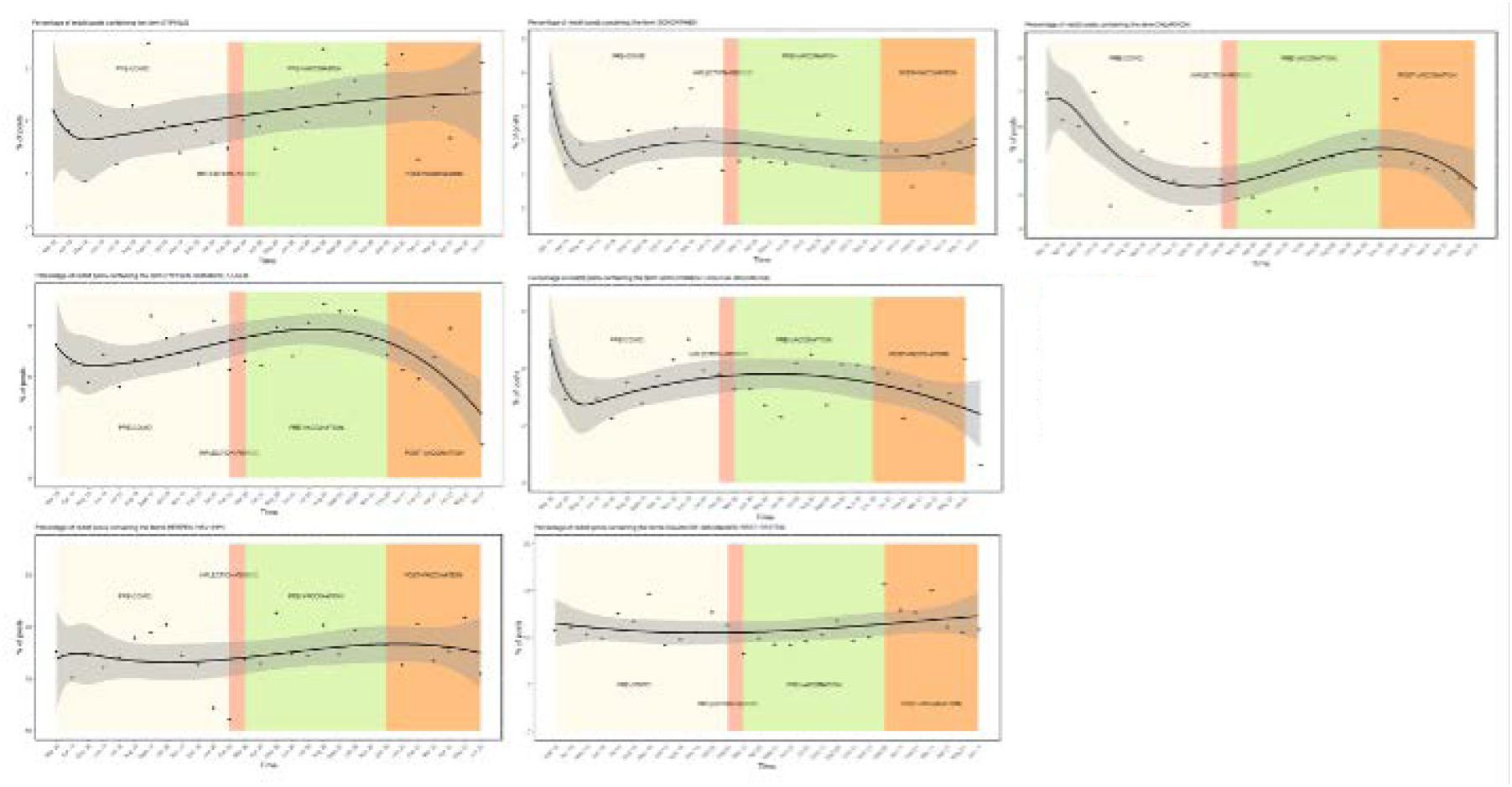
Percentage of Reddit posts containing specific key terms, March 2019-june 2021

## Discussion

### Principal Results

Our study demonstrates that there were changes in the topics posted in STD-related subreddits from pre-COVID through COVID pre-vaccine and COVID post-vaccine periods. The changes in topics discussed likely relate to behavior changes due to COVID related lock-downs, restrictions on in-person gatherings, and closing of non-essential medical services.[20] Regardless of lock-down status, people still engage in sexual behavior (e.g., condomless sex) that will expose them to STDs. However, with reduction of STD testing/treatment these cases are not reflected in surveillance numbers. It is important to understand the sexual health experiences of communities, including symptoms, questions, and behavior patterns, in order to plan for screening and treatment options.

Our results found that “STD risk” as a topic and general “risk” terms as words only appeared in the pre-COVID time period, whereas “HPV” and “warts” only appeared in the COVID pre-vaccine and post-vaccine periods. During the pre-COVID time period, users generated posts related to general STD risk and sexual behavior, seeking advice and support for understanding STD exposure risk for specific sexual behavior or partnership choices. During the two COVID periods this general “STD risk” topic no longer appeared, demonstrating a difference in content moving from a general discussion to specific symptom or etiology-based posts. During the two COVID periods, HPV/warts emerged as a topic. This may be due to increased effort to self-diagnosis symptoms experienced as a result of limited access to diagnostic services. Though reported STD cases declined during the initial lockdown period, 2020 reported cases quickly rebounded and exceed the 2019 case numbers.[3]

Our study’s use of Reddit is a novel way to gain insights into STD symptoms experienced, potential exposures, testing decisions, common questions, and behavior patterns (e.g., during lock down periods). For example, reduction in STD screening may result in observed negative health outcomes due to missed cases, which also impacts onward transmission. The reduction in access to STD testing and treatment during COVID-19 intensified existing barriers to sexual health care, including stigma, judgement, cost, and accessibility.[21] It is important that STD services be maintained, either through telehealth and in-home testing options, or via clinic services with COVID mitigation procedures in place (screening, masking, social distancing).

As Reddit use is anonymous users may discuss sensitive topics with greater detail, more freely than in clinical encounters. The sexual health subreddits had an average volume of unique posts ranging from approximately 700 to 900 per month, thus, Reddit is a frequently used source of information that could guide understanding of behavior, symptoms, and common questions of patients. Sexual health care workers should consider collaboration with Reddit or other social media outlets to leverage the potential benefits of these platforms (anonymous, free, rapid response) while mitigating harm (incorrect diagnoses, faulty recommendations).[22]

## Limitations

Study results should be interpreted considering limitations. LDA is an unsupervised approach with no gold standard to compare to. However, we analyzed the LDA output qualitatively with the use of two independent coders and reached 100% consensus on manual topic labels. As we used posts from an open online forum, we are unable to validate users, however there is little incentive to be dishonest or to post false information on health-related subreddits. Reddit users tend to be younger and are more likely to be male compared to the larger US population, however other demographic trends (e.g., race/ethnicity) mirror the distribution in the US.[23] As men, Black, and Latino people are often underrepresented in STI case data, it is important to gain understanding of their sexual health needs and experiences via alternative data sources.[23] Finally, the precise location of Reddit users are unknown. While we were able to extract posts limited to the United States and in English language, we cannot pinpoint post volume by specific state or local jurisdiction.

## Conclusion

This study demonstrates Reddit has feasibility and utility to enhance understanding of sexual behaviors, STD experiences, and needed health engagement with the public. It is important to prioritize efforts to reduce the spread and impact of STDs through surveillance, screening, and treatment. The COVID-19 pandemic and subsequent stay-at-home orders highlight a critical need for increased access to STD clinics and STD information. Data from anonymous Reddit posts may be leveraged to enhance understanding of the distribution of disease and need for targeted outreach/screening programs.

## Data Availability

All data produced in the present study are available upon reasonable request to the authors

